# Factors Affecting the Reporting of Adverse Event Following Immunization in Ahafo Region of Ghana

**DOI:** 10.64898/2025.12.29.25343182

**Authors:** Sabina Ampon-Wireko, Salaam Dam-Park Laar, Joseph Kwasi Nkrumah

**Author notes:** These authors contributed equally to this work (Sabina Ampon-Wireko, Salaam Dam-Park Laar).

## Abstract

**Background:** Adverse Events Following Immunization (AEFI) remain a significant concern for immunization programs, as underreporting can undermine public confidence and compromise vaccine safety monitoring. Despite significant efforts to address challenges associated with AEFI reporting globally, the AEFI reporting rate in the Ahafo Region of Ghana still remains a challenge. This study investigated healthcare workers’ knowledge and practices regarding AEFI reporting in the Ahafo Region of Ghana, aiming to identify key factors influencing reporting behavior and suggest strategies for improvements for the immunization surveillance system.

**Methods:** A cross-sectional quantitative study was employed in this study to collect data from 1300 frontline health workers who engage in Adverse Events Following immunization surveillance activities. However, the Sample size was determined using Yamane’s formula to obtain a final sample size of 312 with confidence interval of 95% and margin of error being 5%. Multistage sampling approach was employed to ensure that respondents from all six districts in the Ahafo region were proportionately represented. In each district, health facilities were stratified by facility level mainly hospitals, health centers and Community Based Health Planning and Services (CHPS). Descriptive statistics and Poisson regression analysis were used to examine relationships between variables and AEFI reporting practices.

**Results:** The study found that age 26-35 years shows a markedly higher incidence rate ratio (IRR) in all models compared to 18-25 years. Community health nurses and midwives report higher AEFI rates than clinicians; IRR=2.730(1.326-5.228). Respondents with advanced years of work experience reported significantly more AEFI; Model III IRR = 7.938 (95% CI: 2.097-13.008). Only 45.2% of facilities had AEFI reporting forms available, and 40.1% of staff reported no access to reporting guidelines. Training on AEFI significantly increased reporting rate (IRR = 0.485), and lack of access to guidelines reduced it (IRR = 0.814).

**Conclusion:** The study concludes that systemic and individual-level gaps hinder effective AEFI reporting rate. Enhancing access to standard tools, providing regular training, and improving electronic reporting systems are essential. Strengthening caregiver communication and district-level supervision will further improve surveillance and vaccine safety.

## INTRODUCTION

Vaccination is a global health success story, saving millions of lives every year especially from Vaccine Preventable Diseases (VPDs) which accounted for severe to serious morbidities and mortalities mostly among children under 5 years (1). Vaccines reduce risks of getting a disease by working with your body’s natural defenses to build protection (2). Vaccination is an important public health tool that prevents about 2 to 3 million deaths worldwide yearly (1).

Vaccines used in national immunization programmes are extremely safe and effective (3). However, it has been reported that no vaccine is perfectly safe; and adverse reactions may occur since apart from the vaccines themselves, the process of immunization is a potential source of an adverse reaction (4). Together with governments, vaccine manufacturers, scientists and medical experts, WHO’s vaccine safety programme is constantly helping to monitor the safety of vaccines (5).

An Adverse Event Following Immunization (AEFI) is any untoward medical occurrence which follows immunization and which does not necessarily have a causal relationship with the usage of the vaccine (4). The adverse event may be any unfavorable or unintended sign, abnormal laboratory finding, symptom or disease that result following an immunization (6).

Most childhood vaccines have been in use for decades, with millions of people receiving them safely every year, routinely or in response to specific disease threats for example, more than 13 billion doses of COVID-19 vaccines have been safely administered globally since 2021, preventing millions of cases of severe disease and deaths (7).

Before any vaccine is introduced in a country, the vaccine developed in the laboratory undergoes rigorous and stringent testing through multiple phases of clinical trials. Health authorities carefully evaluate the results of these trials to help ensure that the vaccine meets the highest safety and efficacy standards before being considered suitable for use (7).

Most AEFIs are mild, local and systemic, thus, surveillance actions are focused on moderate and severe events. These events are related to several factors, such as the type of vaccine, conditions of administration, storage, and characteristics of the vaccines. Their intensity, however, may vary from mild and expected effects such as local manifestation to moderate and severe events and rare cases, classified as unexpected (8)

Considering the characteristics of the vaccines, children under one year old represent the most AEFI-affected group (8).

Though over 80% of children worldwide are vaccinated, vaccine hesitancy continues to be a public health issue (1). Surveillance on immunization safety is hence crucial to build trust and to reassure the public that AEFIs are being monitored and actions are being taken to reduce risks. This will reduce vaccine-hesitancy and sustain efforts by immunization programs (1).

In Ghana, the Expanded Programme on Immunization (EPI) started in 1978 and since then, several new vaccines have been introduced into the immunization schedule (9). These vaccines are used to prevent Tuberculosis, Poliomyelitis, Diphtheria, Pertussis (Whooping cough), Tetanus, Haemophilus influenza type B, Hepatitis B, Pneumococcal diseases, Rotavirus diarrhoea, Measles, Rubella, Yellow Fever, Neisseria Meningitis and Malaria (9).

The strategies in delivering these vaccines to the beneficiaries are either through routine immunization and supplemental immunization activities or Vaccine Preventable Diseases (VPDs) outbreak reactive campaigns to stop the spread of these diseases in the population (9). Therefore, there are policy directives by Ghana Health Service in collaboration with Food and Drugs Authority (FDA) as to how vaccine safety or Adverse Event Following Immunization can be handled (9).

Empirical studies have shown that AEFIs can be reported by mothers, care givers, health care workers, Community Based Surveillance Volunteers (CBSVs) through the community-based surveillance (CBS) system (6). The EPI programme has outlined two types of AEFI surveillance systems; that is spontaneous and active AEFI surveillance. The spontaneous is where clients or beneficiaries come back to the facility to report on experiences following vaccination. It is a passive way of reporting AEFI after vaccinating a child with a routine vaccine. Mothers or care givers are told to report any abnormal laboratory findings, sign, symptom or a disease to any nearest health facility for medical attention (9).

Active AEFI surveillance on the other hand is where health workers actively look for cases by reviewing records or searching for the AEFI cases at the community level (10). Mostly, Community Health Officers (CHOs) who provide immunization services to children under 5 years weekly in every month in Ghana can actively conduct AEFI case search by doing home visit at the community level to detect AEFI cases following immunization (10). An AEFI when recorded at the facility need to be reported immediately within 24 hours (9). The AEFI investigation team is made up of a clinician, regional EPI Coordinator, Biomedical Laboratory Scientist, regulatory officer from FDA and Deputy Director Public Health.

Investigating these serious AEFI are to enable Technical Advisory Committee (TAC) at national to assign a causality to the AEFI. Adverse Event Following Immunization cases that are not handled well, may prevent care givers from sending their children for subsequent doses (9).

The surveillance flow of AEFI involve detection of cases by the Health Care Worker (HCW), they filled AEFI reporting forms, report is then sent to district EPI Coordinator, District, where it is forwarded to the regional EPI Coordinator (10). At the regional level, the report is sent to the National AEFI focal person.

These AEFI cases are captured into District Health Information Management System (DHIMS2). The assigned data set in DHIM2 are the weekly integrated disease surveillance and response system (Weekly IDSR), monthly integrated disease surveillance and response system (Monthly IDSR) and monthly vaccination report.

The criteria for diagnosing any child or person for AEFI is to use the standard case definition. This include reviewing the records of the beneficiaries to determine when the vaccine was administered. Any ill health reported to health care worker at the health facility level with records showing more than 28 days following vaccination date is not classified as AEFI based on the AEFI surveillance guidelines (11).

The main objective of the AEFI surveillance is to ensure cases are detected early, reported, analyzed and public health actions are taken immediately to foster public confidence in the immunization services (11).

Meanwhile, between the periods of 2020 to 2022 there was no AEFI surveillance flow in the Ahafo region of Ghana. This finding was revealed by PATH Ghana after reviewing DHIMS 2 data. PATH is one of the key partners of health service delivery in Ghana especially in the area of immunization activities with much emphasis on malaria vaccine pilot and expansion in the country. The AEFI case detection rate in the region was 3.5/100,000 live surviving infants in 2023 lower than the suggested 10/100, 000 live surviving infants by the World Health Organization (12). Therefore, immediate steps are needed to ensure well-coordinated AEFI surveillance activities in the Ahafo region, as weak surveillance may compromise access to and utilization of routine immunization activities (13).

It is for this reason that this study investigated healthcare workers’ knowledge and practices regarding AEFI reporting in the Ahafo Region of Ghana to identify key factors influencing reporting behavior with suggested strategies for improvements for the immunization surveillance system.

## MATERIALS AND METHODS

### Study design and population

This study adopted a descriptive cross-sectional design utilizing a quantitative approach. The primary objective was to assess healthcare workers’ knowledge and practices regarding Adverse Events Following Immunization (AEFI) reporting in the Ahafo Region of Ghana. The target population for this study comprised healthcare workers actively engaged in immunization services across the six districts of the Ahafo Region. These included nurses, midwives, public health officers, and other allied health professionals who are expected to identify and report AEFI as part of their responsibilities.

### Sampling Technique and Sample Size

A multistage sampling approach was employed to ensure that respondents from all six districts were adequately represented. The six administrative districts were all included since they encompass the entire Ahafo Region. In the first stage, a list of health facilities that provide immunization services was obtained from the Ghana Health Service Regional Health Directorate according to the districts they belong. From each district, health facilities were stratified by level (hospital, health center, CHPS compound), and a random sample of facilities was drawn proportionally.

In the third stage, within each selected health facility, a list of eligible healthcare workers was generated, and simple random sampling was used to select participants. The number of respondents selected from each district was proportionate to the district’s total number of healthcare workers involved in immunization. This sampling strategy ensured representativeness and minimized selection bias.

The sample size was calculated using Yamane’s formula for the finite population of 312, assuming a 95% confidence interval, a 5% margin of error and immunization workforce of 1300 in the Ahafo Region. The final sample size of 312 was then proportionally distributed among the six districts based on their healthcare workforce size involved in immunization activities.

A structured, self-administered questionnaire was used to collect data from study participants. The questionnaire was developed based on guidelines from the World Health Organization (WHO), the Ghana Health Service AEFI reporting protocol, and existing literature on immunization and vaccine safety surveillance. The tool comprised both closed-ended items covering the following domains: Demographic and professional characteristics (e.g., age, gender, years of experience, professional role), Knowledge of AEFI (e.g., definitions, types, symptoms, reporting protocols), Practices related to AEFI reporting (e.g., methods of reporting, use of forms, informing caregivers), Facility-level factors (e.g., availability of guidelines, training opportunities, workload)

### Data Analysis

Completed questionnaires were manually reviewed for completeness before data entry. Data were entered into Stata version 17.0 and cleaned to address inconsistencies and missing values. Descriptive statistics (frequencies, percentages, means, and standard deviations) were computed to summarize demographic characteristics, knowledge levels, and reporting practices. For inferential analysis, Poisson regression with robust standard errors was used to identify factors associated with AEFI reporting. This analytical approach was appropriate given the count nature of the outcome variable (number of AEFI cases reported). Incidence Rate Ratios (IRRs) and 95% confidence intervals were reported. Variables that were significant in the bivariate analysis at p < 0.10 were included in the multivariate Poisson regression model. Missing data were handled using listwise deletion for cases with excessive missing values (>20% of items unanswered). However, for minor missingness (<5%), mean imputation was applied for scale-based variables to preserve sample size and statistical power.

### Ethical Considerations

Ethical clearance for the study was obtained from the Catholic University of Ghana Institutional Research Board, which ensures adherence to ethical principles in research involving human subjects (Ethical Review number CUG/ERB/25-109). In addition, permission to conduct the study was obtained from the Ahafo Regional Health Directorate and the various District Health Directorates. Written informed consent was sought from each participant after a thorough explanation of the study’s purpose, procedures, and their rights, including voluntary participation and confidentiality. Participants were assured that their responses would be used solely for academic purposes and would not affect their employment or relationships within the health system. All data were anonymized during analysis, and access was restricted to the research team.

## RESULTS

### Demographic data

Majority of the respondents were females (69.2%), and those aged between 26-35 years (82.6%), as well as Christians (93.0%). Most respondents were Community Health Nurses (60.3%) with less than 5 years of working experience (73.7%). The marital status of respondents was predominantly single (53.2%) or married (45.5%). The respondents were distributed across various healthcare facilities, including Health Centers (40.1%) and Hospitals (45.5%), with a smaller proportion working in CHPS (14.4%). This is shown in Table 1.

**Table 1:** Demographic characteristics of study participants.

| Variable | Frequency<br>(n=312) | Percent (%) |
| --- | --- | --- |
| **Sex** |  |  |
| Male | 96 | 30.8 |
| Female | 216 | 69.2 |
| **Age Grouping** |  |  |
| 18 – 25 | 12 | 3.9 |
| 26 – 35 | 258 | 82.6 |
| 36 – 45 | 38 | 12.2 |
| 45 + | 4 | 1.3 |
| **Religion** |  |  |
| Christian | 290 | 93.0 |
| Muslim | 20 | 6.4 |
| African Trad. Religion | 2 | 0.6 |
| **Cadre of respondents** |  |  |
| Clinician | 14 | 4.5 |
| Community Health Nurse | 188 | 60.3 |
| Disease Control Officer | 38 | 12.2 |
| Field Technician (DC) | 10 | 3.2 |
| Midwife | 62 | 19.8 |
| **Working Experience of respondents** |  |  |
| < 5year | 230 | 73.7 |
| 5-10 years | 50 | 16.0 |
| 10+years | 32 | 10.3 |
| **Marital status** |  |  |
| Single | 166 | 53.2 |
| Married | 142 | 45.5 |
| Divorced | 4 | 1.3 |
| **Type of facility** |  |  |
| Hospital | 142 | 45.5 |
| Health Center | 125 | 40.1 |
| CHPS | 45 | 14.4 |

**Source:** *Field Survey, 2025*

### Number of reported cases of AEFI by districts in the Ahafo Region

Figure 1 illustrates the spatial distribution of Adverse Events Following Immunization (AEFI) cases reported across districts in Ahafo region of Ghana. The map categorizes districts based on reported AEFI cases, with four districts (Asutifi North, Asunafo North Municipal, Asutifi South, and Tano South Municipal) reporting moderate cases (5-10) and two districts (Tano North Municipal and Asunafo South) reporting fewer cases (<5) with no district reporting cases up to the threshold of AEFI reports suggested by the Ghana Health Service.

**Figure 1:**
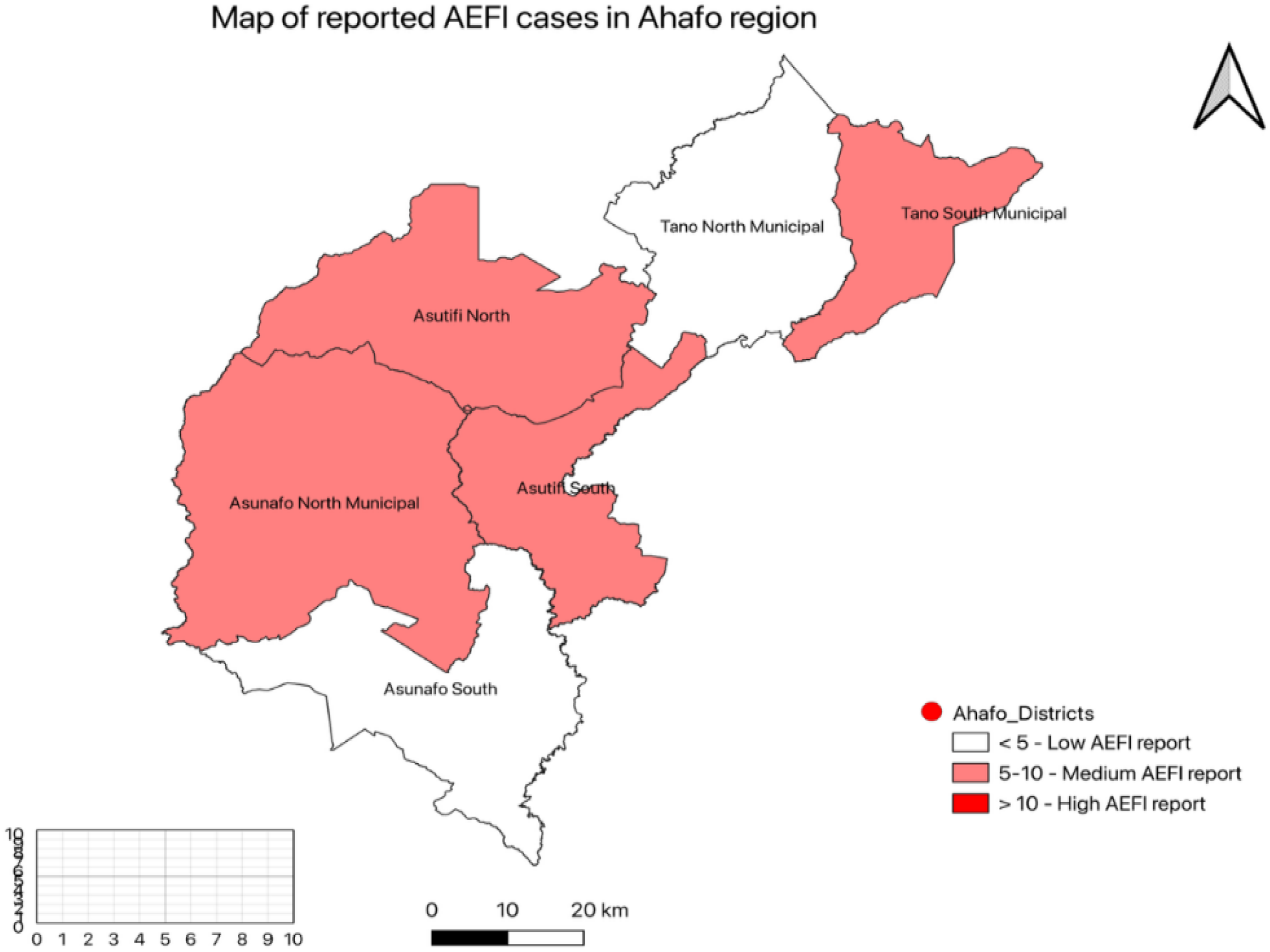
Reported cases of AEFI according to Districts in Ahafo region. **Source:** *Field Survey, 2025*

### Knowledge of healthcare workers and facility practice regarding Adverse Events Following Immunization (AEFI) reporting

The data indicates that while there is a high level of awareness and presence of AEFI surveillance guidelines in the healthcare facilities (93.6%), there are gaps in the standardization of reporting forms where 54.8% lack standard forms for reporting in their facilities, accessibility of guidelines to all staff (40.1%) not accessible, and training on AEFI not trained in 42.9% of respondents. The primary method of reporting AEFI cases is through an electronic reporting system (69.2%), which is a positive indicator for efficient data collection and analysis. However, the dissemination of guidelines is not uniformly known or practiced, with a significant proportion (46.8%) of respondents being unaware of how guidelines are disseminated **(Table 2)**.

**Table 2:** Knowledge and facility practice on AEFI reporting.

| Variable | Frequency | Percent (%) |
| --- | --- | --- |
| Reporting AEFI cases to the next level in your facility |  |  |
| By filling Paper based forms | 292 | 93.6 |
| Electronic reporting system | 4 | 1.3 |
| Telephone call | 16 | 5.1 |
| Have Standard AEFI reporting forms in this health facility |  |  |
| Yes | 141 | 45.2 |
| No | 171 | 54.8 |
| Have AEFI surveillance guidelines in this health facility |  |  |
| Yes | 291 | 93.3 |
| No | 21 | 6.7 |
| The Guidelines include information on how to report AEFI |  |  |
| Yes | 187 | 59.9 |
| No | 125 | 40.1 |
| The AEFI surveillance guidelines accessible to all healthcare staff |  |  |
| Yes | 187 | 59.9 |
| No | 125 | 40.1 |
| The guidelines disseminated to the staff |  |  |
| Printed copies | 104 | 33.3 |
| Soft copies | 62 | 19.9 |
| Don't know | 146 | 46.8 |
| Received training of any form or format relating to AEFI |  |  |
| Yes | 178 | 57.1 |
| No | 134 | 42.9 |

**Source:** *Field Survey, 2025*

### Vaccine handling and AEFI reporting practices

**Table 3** presents responses related to vaccine handling and AEFI reporting practices. The data suggests that while most respondents follow proper vaccine handling procedures such as checking temperature (93.6%), checking expiry dates (76.3%), and using VVMs (95.2%), there are areas of concern, such as informing caregivers about reporting adverse events, where only 46.8% of respondents do so. Additionally, a significant majority (71.8%) find the AEFI reporting process time-consuming, which could potentially impact reporting rates.

**Table 3:** Vaccine handling and AEFI reporting practices.

| **Variable** | **Frequency** | **Percent (%)** |
| --- | --- | --- |
| Check vaccines temperature range of +2 to + 8 degree Celsius |  |  |
| Yes | 292 | 93.6 |
| No | 20 | 6.4 |
| Check for shelf life or expiry date of the vaccines before its usage |  |  |
| Yes | 238 | 76.3 |
| No | 74 | 23.7 |
| Read your vaccine vial monitor (VVM) before its administration |  |  |
| Yes | 297 | 95.2 |
| No | 15 | 4.8 |
| Condition your icepacks before putting vaccines them in the vaccine carrier |  |  |
| Yes | 312 | 100.0 |
| No | 0 | 0.0 |
| Perform shake test on suspected freeze sensitive frozen vaccines |  |  |
| Yes | 231 | 74.0 |
| No | 81 | 26.0 |
| Inform care givers to report any adverse events to the nearest health facility |  |  |
| Yes | 146 | 46.8 |
| No | 166 | 53.2 |
| Find AEFI reporting process to be time consuming |  |  |
| Yes | 224 | 71.8 |
| No | 88 | 28.2 |
| Acknowledge reporting AEFI to the next level as part of your work duties |  |  |
| Yes | 268 | 85.9 |
| No | 44 | 14.1 |

**Source:** *Field Survey, 2025*

### Socio-demographic factors influencing AEFI reporting among healthcare workers

Table 4 presents the results of a Poisson regression model examining the influence of socio- demographic factors on the count of AEFI reported. It was indicated that individuals aged 26-35 years have a significantly higher incidence rate ratio (IRR) across all models, indicating a strong association with AEFI reporting. Model III shows an IRR of 17.507 (8.201-37.210), suggesting that this age group reports AEFI at a rate approximately 17.5 times higher than the reference group (18-25 years). Respondents’ religion also showed that Muslims have a significantly higher IRR compared to Christians across all models. In Model III, the IRR is 10.251 (1.655-63.522), indicating that Muslims report AEFI at a rate about 10 times higher than Christians.

**Table 4:** Poisson regression model indicating influence of socio-demographic factors on count of AEFI reported.

| Variable | Model I<br>Crude IRR<br>[95%CI] | Model II<br>Adjusted IRR<br>[95%CI] | Model III<br>Adjusted IRR<br>[95%CI] |
| --- | --- | --- | --- |
| **Sex** |  |  |  |
| Male | 1 | 1 | 1 |
| Female | 2.667 (0.607-11.713) | 2.109 (0.656-12.197) | 2.553 (0.687-9.491) |
| **Age grouping** |  |  |  |
| 18 – 25 | 1 | 1 | 1 |
| 26 – 35 | 6.789 (1.981-2.331)* | 10.655 (3.226-35.188)* | 17.507 (8.201-37.210)* |
| 36 – 45 | 1.260 (0.342-4.887) | 1.821 (1.136-4.581)* | 5.287 (4.044-6.901)* |
| 45 + | 1.226 (0.897-5.567) | 1.015 (0.177-5.818) | 5.361 (1.761-6.416)* |
| **Religion** |  |  |  |
| Christian | 1 | 1 | 1 |
| Muslim | 3.954 (1.197-13.068)* | 6.844 (1.867-25.092)* | 10.251 (1.655-63.522)* |
| African Trad. Religion | 0.173 (0.038-0.870)* | 0.016 (0.001-0.673)* | 0.140 (0.026-0.744)* |
| **Cadre of respondents** |  |  |  |
| Clinician | 1 | 1 | 1 |
| Community Health<br>Nurse | 5.331 (2.307-23.071)* | 8.046 (5.249-16.752)* | 2.730 (1.326-5.228)* |
| Disease Control Officer | 1.002 (0.541-1.847) | 1.081 (0.637-1.834) | 2.948 (0.229-38.001) |
| Field Technician (DC) | 1.201 (0.362-2.764) | 0.995 (0.503-1.971) | 0.699 (0.074-6.643) |
| Midwife | 4.413 (1.204-16.207)* | 5.914 (2.454-7.646)* | 1.307 (1.142-4.208)* |
| **Working experience of**<br>**respondents** |  |  |  |
| < 5year | 1 | 1 | 1 |
| 5-10 years | 1.255 (0.363-4.341) | 1.478 (0.421-5.187) | 0.799 (0.171-3.739) |
| 10+years | 1.540 (1.021-7.869)* | 3.907 (1.877-8.117)* | 7.938 (2.097-13.008)* |
| **Marital status** |  |  |  |
| Single | 1 | 1 | 1 |
| Married | 2.104 (0.721-6.145) | 2.119 (0.683-6.577) | 1.374 (0.434-4.347) |
| Divorced | 3.249 (2.750-12.008)* | 0.339 (0.118-0.897)* | 0.838 (0.129-0.981)* |
| **Type of facility** |  |  |  |
| CHPS | 1 | 1 | 1 |
| Health Center | 0.450 (0.126-1.605) | - | - |
| Hospital | 0.396 (0.111-1.415) | - | - |

**Source:** *Field Survey, 2025*

Again, Community Health Nurses and Midwives have a significantly higher IRR in Models I and II, but the IRR decreases in Model III. In Model III, the IRR is 2.730 (1.326-5.228) and 1.307 (1.142-4.208), suggesting that Community Health Nurses and Midwives report AEFI at a rate about 2.7 and 1.3 times higher than Clinicians respectively.

Likewise, respondents with 10+ years of experience have a significantly higher IRR in Model III (AIRR=7.938; 95%CI: 2.097-13.008), indicates that those with 10+ years of experience report AEFI at a rate about 7.9 times higher than those with less than 5 years of experience.

Lastly, The IRR for divorced individuals varies significantly across models, with Model III reporting a decreased IRR of 0.838 (0.129-0.981), suggesting that after adjustments, being divorced is associated with a lower rate of AEFI reporting compared to being single.

### Influence of staff knowledge and facility practices on count of AEFI reported

**Table 5** lists various variables related to AEFI reporting practices and their association with the count of AEFI reported. Model III provides the most adjusted estimates with significant findings including reporting AEFI cases via an electronic reporting system is associated with a lower IRR (AIRR=0.690; 95%CI: 0.151-0.922) compared to using paper-based forms, indicating a reduction in AEFI reporting.

**Table 5:** Poisson regression model indicating influence of knowledge and facility practices on count of AEFI reported.

| Variable | Model I | Model II | Model III |
| --- | --- | --- | --- |
|  | Crude IRR | Adjusted IRR | Adjusted IRR |
|  | [95%CI] | [95%CI] | [95%CI] |
| **Reporting AEFI cases to the next level in your facility** |  |  |  |
| By filling Paper based forms | 1 | 1 | 1 |
| Electronic reporting system | 0.293 (0.096-0.893)* | 0.257 (0.066-0.410)* | 0.690 (0.151-0.922)* |
| Telephone call | 1.403 (0.195-10.108) | 1.298 (0.188-8.946) | 1.280 (0.217-7.546) |
| **Have standard AEFI reporting forms in this health facility** |  |  |  |
| Yes | 1 | 1 | 1 |
| No | 1.099 (0.389-3.099) | 1.158 (0.397-3.378) | 1.096 (0.357-3.367) |
| **Have AEFI surveillance guidelines in this health facility** |  |  |  |
| Yes | 1 | 1 | 1 |
| No | 1.066 (0.146-7.785)* | 1.238 (0.145-10.565)* | 0.814 (0.362-0.907)* |
| **The guidelines include information on how to report AEFI** |  |  |  |
| Yes | 1 | 1 | 1 |
| No | 1.122 (0.398-3.161) | 1.185 (0.427-3.286) | 1.038 (0.104-10.361) |
| **The AEFI surveillance guidelines accessible to all healthcare staff** |  |  |  |
| Yes | 1 | 1 | 1 |
| No | 1.122 (0.398-3.161) | 1.185 (0.427-3.286) | 1.038 (0.104-10.361) |
| **The guidelines disseminated to the staff** |  |  |  |
| Printed copies | 1 | 1 | 1 |
| Soft copies | 3.354 (0.631-17.833) | 3.049 (0.570-16.295) | 2.790 (0.426-18.258) |
| Don't know | 2.849 (0.616-13.177) | 2.6078 (0.586-11.597) |  |
| **Received training of any form or format relating to AEFI** |  |  |  |
| Yes | 1 | 1 | 1 |
| No | 0.738 (0.253-2.155) | 0.765 (0.263-2.225) | 0.485 (0.206-0.614)* |
| **Facility type** |  |  |  |
| CHPS | 1 | - | - |
| Health Center | 0.450 (0.126-1.605) | - | - |
| Hospital | 0.396 (0.111-1.415) | - | - |
| **Condition of vaccines in facility for use?** |  |  |  |
| Very often | 1 | 1 | - |
| Not often | 1.532 (0.903-3.451) | 1.655 (0.995-3.152) | - |

**Source:** *Field Survey, 2025*

Again, facilities without AEFI surveillance guidelines have a reduced rate of reporting AEFI cases IRR (AIRR=0.814; 95%CI: 0.362-0.907) compared to those with guidelines.

Receiving training on AEFI is associated with a higher IRR (since "No" training has an IRR of 0.485, implying that not receiving training reduces AEFI reporting).

### The relationship between staff practices, conditioning, and AEFI reporting in immunization settings

**Table 6** shows the results of three Poisson regression models assessing how staff vaccine- handling practices demographic and facility characteristics influence the count of adverse events following immunization (AEFI) reported.

**Table 6:** Poisson regression model indicating influence of staff practices and conditioning on count of AEFI reported.

| Variable | Model I | Model II | Model III |
| --- | --- | --- | --- |
|  | Crude IRR [95%CI] | Adjusted IRR [95%CI] | Adjusted IRR [95%CI] |
| Check vaccines temperature range of +2 to + 8 degree Celsius |  |  |  |
| Yes | 1 | 1 | 1 |
| No | 1.123 (0.154-8.183) | 1.414 (0.183-10.903) | 1.030 (0.138-7.679) |
| **Check for shelf life or expiry date of the vaccines before its usage** |  |  |  |
| Yes | 1 | 1 | 1 |
| No | 0.877 (0.251-3.067) | 0.956 (0.270-3.390) | 1.149 (0.341-3.875) |
| **Read your vaccine vial monitor (VVM) before its administration?** |  |  |  |
| Yes | 1 | 1 | 1 |
| No | 1.523 (0.212-10.920) | 1.426 (0.177-11.505) | 1.057 (0.119-9.361) |
| **Condition your icepacks before putting vaccines them in the vaccine carrier** |  |  |  |
| Yes | 1 | 1 | 1 |
| No | 1.132 (0.339-14.896) | 1.107 (0.330-17.896) | 0.987 (0.436-16.788) |
| **Perform shake test on suspected freeze sensitive frozen vaccines** |  |  |  |
| Yes | 1 | 1 | 1 |
| No | 2.897 (0.458-12.122) | 2.587 (0.556-11.909) | 2.234 (0.515-11.563) |
| **Inform care givers to report any adverse events to the nearest health facility** |  |  |  |
| Yes | 1 | 1 | 1 |
| No | 3.2249 (0.916-11.358) | 2.874 (0.813-10.166) | 2.910 (0.850-9.961) |
| Don't know |  |  |  |
| **Find AEFI reporting process to be time consuming** |  |  |  |
| Yes | 1 | 1 | 1 |
| No | 0.678 (0.204-5.543) | 0.598 (0.1890-4.981) | 0.510 (0.172-2.054) |
| **Age Grouping** |  |  |  |
| 18-25 | 1 | 1 | - |
| 26-35 | 6.789 (1.981-2.331)* | 10.655 (3.382-33.564)* | - |
| 36-45 | 1.260 (0.342-4.887) | 1.821 (1.338-7.387)* | - |
| 45+ | 1.226 (0.897-5.567) | 1015 (0.185-5.586) | - |
| **Facility type** |  |  |  |
| CHPS | 1 | - | - |
| Health Center | 0.450 (0.126-1.605) | - | - |
| Hospital | 0.396 (0.111-1.415) | - | - |

Across all three models, none of the vaccine management practices including checking cold-chain temperature, confirming expiry dates, reading VVM status, conditioning icepacks, performing shake tests, or instructing caregivers to report AEFIs showed a statistically significant association with AEFI reporting counts. Their incidence rate ratios (IRRs) had wide confidence intervals overlapping 1, indicating no meaningful effect after adjusting for covariates.

However, age showed a strong and significant influence in the adjusted models. Staff aged 26– 35 years had significantly higher AEFI reporting rates compared with the 18–25 year group in both Model I and Model II, suggesting that the more experienced tend to report more AEFIs. The age category 36–45 years also showed a significant increase in reporting rates in Model II. Facility type and perceptions of whether the reporting process is time-consuming did not significantly predict AEFI reporting.

## DISCUSSION

### Socio-demographic factors influencing AEFI reporting among healthcare workers

Although female healthcare workers showed higher incidence rate ratios (IRRs) of AEFI reporting across all models, the associations were not statistically significant. This finding is consistent with a study in Ethiopia, which found no significant difference between male and female healthcare workers in AEFI reporting (14). However, it contradicts a study in India, where female health workers reported more frequently due to increased involvement in routine immunization services (15). The non-significance in this study may suggest that both male and female staff are equally sensitized to AEFI reporting procedures in the Ahafo Region.

In the current study, age was a strong and significant predictor of AEFI reporting. Healthcare workers aged 26–35 years had significantly higher reporting rates than those aged 18–25 years. This trend remained consistent across all models. The adjusted IRR in Model III was particularly high (17.507; 95% CI: 8.201–37.210), indicating that younger middle-aged professionals may be more vigilant or better trained. This aligns with research from Nigeria and Kenya, where younger mid-career healthcare workers were more active in surveillance-related activities (16). One possible explanation is that younger professionals may have more recent training or exposure to updated guidelines on AEFI reporting.

Religious affiliation was also significantly associated with AEFI reporting. Muslim respondents had significantly higher IRRs compared to Christians (Model III IRR = 10.251; 95% CI: 1.655–63.522), while those practicing African Traditional Religion were significantly less likely to report (Model III IRR = 0.140; 95% CI: 0.026–0.744). The higher reporting among Muslims could reflect religious community engagement in health initiatives, as found in studies in Northern Ghana and parts of Nigeria where Muslim community leaders actively support immunization campaigns (17). The lower rates among traditional religion adherents may point to mistrust in modern medicine or limited involvement in structured health systems, as noted in earlier studies

(18). This divergence underscores the need for culturally sensitive AEFI communication strategies.

The professional cadre of healthcare workers significantly influenced AEFI reporting. Community Health Nurses (CHNs) and Midwives had significantly higher adjusted IRRs compared to clinicians. This may be attributed to their frontline role in vaccine administration, which increases their exposure to potential AEFIs. Previous studies in Ghana and Uganda reported similar trends, with CHNs more likely to detect and report AEFIs due to their routine contact with clients during outreach and static services (19,20). Conversely, Disease Control Officers and Field Technicians did not show significant associations, possibly because they play more supervisory or supportive roles and may not directly encounter post-vaccination clients.

Healthcare workers with over 10 years of experience were significantly more likely to report AEFIs (Model III IRR = 7.938; 95% CI: 2.097–13.008). This finding is in line with studies from South Africa and Zambia, where more experienced professionals demonstrated greater familiarity with AEFI reporting protocols and the confidence to report (19). Interestingly, those with 5–10 years of experience did not show significant differences, suggesting that experience beyond a decade may be necessary for full integration of AEFI reporting into practice.

Divorced respondents had significantly lower IRRs in the adjusted models, reversing the strong positive association seen in the crude model. This suggests confounding factors may have influenced the initial association. There is limited literature linking marital status directly with AEFI reporting. However, stress and social support differences among divorced individuals may indirectly affect professional performance and vigilance in reporting, as noted in occupational health studies (21).

The type of health facility (CHPS, Health Centre, or Hospital) was not a significant predictor of AEFI reporting and was excluded from the adjusted models. This contrasts with findings from a study in Kenya where hospital-based staff reported fewer AEFIs, possibly due to heavy workloads and limited time (22). The lack of significance in this study may suggest relatively uniform training and reporting structures across different facility types within the Ahafo Region.

### Influence of knowledge and facility practices on count of AEFI cases reported

In this current study, it was noted that Healthcare workers using electronic reporting systems had significantly lower incidence rate ratios (IRRs) of AEFI reporting compared to those using paper-based forms.

This finding may appear counterintuitive, as electronic systems are often assumed to improve reporting efficiency. However, similar observations have been made in low-resource settings where digital infrastructure is weak, internet connectivity is unreliable, and staff are insufficiently trained in using electronic platforms (23). In contrast, paper-based systems remain more familiar and accessible to staff at lower-level facilities in Ghana (24). This suggests that unless electronic systems are fully functional and user-friendly, they may hinder rather than enhance AEFI reporting.

Additionally, with reference to the availability of AEFI reporting forms he presence of standard AEFI reporting forms in a facility was not significantly associated with reporting in any model. This suggests that merely having the forms available does not guarantee usage or effective reporting. This aligns with previous findings in sub-Saharan Africa, where underreporting persisted despite the availability of surveillance tools due to lack of motivation or follow-up (17). This emphasizes the importance of guidelines as a reference point for health workers. The presence of guidelines enhances clarity on what to report, how to report it, and who to notify, as demonstrated in studies from Uganda and Tanzania (5,12). The mode of disseminating surveillance guidelines (printed copies, soft copies, unknown) did not significantly influence AEFI reporting in adjusted models. However, those who received soft copies showed a trend toward higher reporting (Model III IRR = 2.790; 95% CI: 0.426–18.258), though this was not statistically significant due to wide confidence intervals likely a result of small subgroup sizes. Previous studies suggest printed materials remain more effective in settings with limited digital access (24), though the evidence is mixed.

Notably, healthcare workers who had not received any training on AEFI had significantly lower rates of reporting in Model III (IRR = 0.485; 95% CI: 0.206–0.614). This underscores the critical role of training in building staff capacity and confidence to detect and report adverse events. Similar findings have been reported in multiple contexts, including Kenya and Nigeria, where training improved both the quantity and quality of AEFI reports (12,22). This result strongly supports the need for periodic in-service training, refresher courses, and mentorship in Ghana’s immunization program.

The study explored how vaccine handling and caregiver engagement practices affect AEFI reporting. Surprisingly, HCWs who did not check the vaccine temperature range were associated with a slightly higher likelihood of reporting AEFIs, although this was not statistically significant (IRR = 1.030, 95% CI: 0.138–7.679). This may reflect increased reporting due to poor handling practices that result in more AEFIs, rather than deliberate surveillance.

Furthermore, not informing caregivers to report potential adverse events was associated with higher AEFI reporting (IRR = 2.910, 95% CI: 0.850–9.961). This suggests that in the absence of caregiver feedback, healthcare workers may rely more on clinical suspicion and internal facility-based surveillance mechanisms. However, the wide confidence interval indicates potential variability, and further research may be needed.

### Relationship between staff practices, conditioning, and AEFI reporting in immunization settings

The Poisson regression model examined the association between various healthcare worker characteristics and the number of AEFI reports submitted within health facilities. The model was statistically significant, indicating that the set of predictors collectively contributed to variations in the reporting of adverse events. This finding aligns with similar studies that underscore the influence of both individual and institutional factors on AEFI reporting behavior (18,24).

One of the most salient findings from the analysis was that healthcare workers who had received prior training on AEFI surveillance were significantly more likely to report adverse events than their untrained counterparts. Specifically, trained personnel were 1.8 times more likely to submit AEFI reports, even after controlling for other factors. This is consistent with findings by Putri and colleagues, who reported that training interventions led to a notable increase in pharmacovigilance reporting rates in Nigeria (25). Similarly, a study conducted in Kenya found that AEFI training improved both the knowledge and willingness of healthcare providers to report adverse events, suggesting that capacity-building plays a critical role in enhancing vaccine safety monitoring systems (22).

In addition, the analysis revealed that years of professional experience had a modest but significant negative association with AEFI reporting. For every additional year of work experience, there was a 5% reduction in the likelihood of reporting an adverse event. This may reflect reporting fatigue or overconfidence among more experienced workers, who might underappreciate the importance of documenting seemingly minor post-immunization reactions. Previous studies have observed similar trends, where younger or less experienced healthcare workers tend to adhere more strictly to reporting guidelines due to recent exposure to training or supervision (21). This finding highlights the importance of continuous refresher training regardless of years of service.

Contrary to expectations, the type of facility (public vs. private) and the cadre of the health worker (nurse vs. physician) were not significantly associated with the number of AEFI reports submitted. These findings differ from a study in Uganda, where public sector facilities reported more events due to better integration with national surveillance systems (26). The lack of association in the present study could suggest a relatively uniform implementation of reporting procedures across facility types in the study area, or possibly underreporting due to systemic barriers not captured in this model, such as workload, supervision, or access to reporting tools.

The overall model fit indicated no serious violation of the equidispersion assumption, suggesting that Poisson regression was appropriate for the count nature of the outcome variable. However, future research may consider alternative modeling techniques such as negative binomial regression, especially if overdispersion is detected in similar datasets.

In summary, the results affirm the pivotal role of AEFI training in enhancing surveillance and reporting practices among healthcare workers. They also suggest that experience alone may not be a sufficient driver of reporting behavior without continuous professional development. These findings support the broader global health agenda that emphasizes strengthening pharmacovigilance systems as part of sustainable immunization programs.

## CONCLUSION

This study provides important information about the factors affecting adverse event following immunization reporting among health workers. It analyses data collected among the health care workers to determine the availability of adverse immunization guidelines and tools, immunization practices influencing AEFI occurrence and demographic factors affecting AEFI reporting.

The findings indicated that availability of adverse event following immunization reporting forms, guidelines and communication leaflets are very essential materials needed to enhance the AEFI reporting within the surveillance sites in the region. This study also revealed that advocacy and communication on the need to educate care givers and vaccine beneficiaries about reporting adverse event following immunization be it sign, symptom, any laboratory findings or a disease to the nearest health care provider was not at it optimum.

Looking at the observed gaps in knowledge, limited accessibility to AEFI surveillance guidelines, and variations in reporting practices, it is important to propose practical, evidence-based interventions that build the capacity of healthcare workers and strengthen the existing immunization surveillance system. Furthermore, the results underscore the importance of structured training, the availability of standardized reporting tools, and the role of facility-level protocols in improving AEFI reporting outcomes.

## Data Availability

The study employed Primary data"

